# Ovarian Hyperstimulation Syndrome: The STOP-OHSS RCT of outpatient paracentesis and survey on preventive practices

**DOI:** 10.1101/2025.07.18.25331764

**Authors:** Mostafa Metwally, Katie Ridsdale, Munya Dimairo, Clare Pye, Ying Cheong, Lauren Desoysa, Andrew Drakely, Isaac Evbuomwan, Jane Hughes, Amanda Loban, Raj Mathur, Kirsty McKendrick, Cara Mooney, Rich Simmonds, Liz Taylor, Siqi Wu, David White, the STOP-OHSS Study Group

## Abstract

**Introduction:** Ovarian Hyperstimulation Syndrome (OHSS) is a potentially serious complication of ovarian stimulation and Assisted Reproductive Technologies. This study aimed to assess the effectiveness of early outpatient paracentesis in preventing symptom progression and hospitalisation of OHSS patients, and resulting in earlier resolution of the condition.

**Methods:** A randomised controlled trial was conducted, randomly allocating fertility centre patients with moderate or severe OHSS to either conservative management (n = 5), or outpatient paracentesis with increased monitoring (n = 3). Randomisation was computer-generated, and the primary outcome was OHSS-related hospitalisation. Due to recruitment challenges, particularly pertaining to the SARS-CoV-2 (COVID-19) pandemic, the trial was closed early after randomising eight participants from three sites. A post-trial survey was conducted to examine changes in clinical practices at UK fertility centres during and after the COVID-19 pandemic.

**Results:** Of the eight participants, two were hospitalised for OHSS-related reasons. Six participants’ symptoms resolved during follow-up, while two had unknown outcomes. No significant adverse events related to paracentesis were reported. The post-trial survey, completed by 16 fertility centres, revealed that 31% of respondents believed OHSS incidence had decreased during and after COVID-19. Many centres reported increased use of preventive strategies during and after COVID-19. For example, 91% of sites reported that they increased their elective freeze rates; 53% increased their use of the antagonist protocol; and 53% increased their use of Gonadotropin-Releasing Hormone (GnRH) trigger for final oocyte maturation.

**Conclusions:** The small sample size precludes definitive conclusions, however, outpatient paracentesis appears to be a safe option for managing OHSS. The study highlights challenges in researching rare conditions with rapid disease progression. The survey demonstrated that these challenges may also be reflective of radical changes in practice during the pandemic, which may have decreased the risk of OHSS significantly.

The trial registration number was ISRCTN71978064, and the project was funded by the National Institute for Health and Care Research Health Technology Assessment programme (NIHR128137).

## ARTICLE SUMMARY

### Strengths and limitations of this study

- Contributes to the growing body of evidence supporting the safety and acceptability of paracentesis;
- Provides novel insights into the evolution of site practices during the COVID-19 pandemic
- Premature trial termination due to recruitment challenges, precluding definitive conclusions
- Challenges in outcome adjudication, potentially affecting data interpretation

## INTRODUCTION

Ovarian Hyperstimulation Syndrome (OHSS) is a common complication of Assisted Reproductive Technology, causing fluid accumulation and discomfort. OHSS can be classified as Mild, Moderate, Severe, or Critical. In its mild form, OHSS is relatively common and may be considered a typical reaction to ovarian stimulation procedures. As the condition progresses to moderate severity, patients often experience symptoms related to fluid accumulation in the extra-vascular spaces, such as ascites and abdominal swelling which together with ovarian enlargement can lead to significant discomfort. Severe cases are characterised by significant fluid retention and dehydration, which can alter blood composition, and in critical instances, OHSS may cause pleural effusion, deep vein thrombosis, pulmonary embolisms, breathing difficulties, multi-organ dysfunction, and in rare cases may even be fatal. Current treatment primarily involves monitoring, with intervention typically only occurring once the disease reaches a severe state. This then requires hospitalisation and inpatient management, usually including inpatient paracentesis, in which the ascitic fluid is drained. Moderate to severe OHSS affects up to 8% of In Vitro Fertilisation patients ^1^.

OHSS can also be categorised based on its onset time. Early-onset OHSS typically occurs within a week of receiving the final human chorionic gonadotropin (hCG) injection and is usually attributed to the ovarian stimulation medications used during treatment. Late-onset OHSS, on the other hand, generally develops 10 or more days after hCG administration and is triggered by the body’s own hCG production in the event of pregnancy.

Some evidence suggests early outpatient paracentesis may prevent hospitalisation and be more cost-effective than conservative management^2–8^. However, robust randomised controlled trials are lacking. The STOP-OHSS study (Shaping and Trialling Outpatient Protocols for Ovarian HyperStimulation Syndrome) aimed to provide stronger evidence on whether early outpatient paracentesis could safely prevent symptom escalation and hospitalisation in OHSS patients. The study planned to assess the clinical and cost-effectiveness, and safety and acceptability of this procedure.

However, the trial struggled to recruit sufficient participants and as a result was closed early after randomisation of eight participants. Discontinuation of trials results in considerable financial implications, due to the time and money invested in the design and set-up of trials^9^. The SARS-CoV-2 (COVID-19) pandemic presented unique challenges for clinical trials. The implemented restrictions, lockdowns, redeployment of clinical staff, and social distancing, had implications on trial units, impacting the participating centres and the recruitment of participants. Mitchel et al (2020)^10^ have described a number of problems affecting research trials caused by the pandemic, on: trial recruitment and consent; communication with site staff and other stakeholders; intervention delivery; data collection; re-starting; and training. The STOP-OHSS study was affected by the impact of COVID-19 on the capacity of staff at sites, which hindered the ability to open new sites and recruit participants. However, this may have not been the only factor hindering recruitment.

Though OHSS is a common side effect of Assisted Reproductive Technologies, severe OHSS is still a relatively rare syndrome; between 38-103 severe cases have been reported per year since 2015^11^. During our study, centres reported that they were seeing fewer potentially eligible participants. We hypothesised that one factor contributing to this recruitment challenge could be a reduced incidence of OHSS compared to before the COVID-19 pandemic. This was suggested by data collected in the STOP-OHSS qualitative sub-study conducted in 2020-2021^12^, as well as anecdotal evidence from sites which implied that more preventive practices were being used to prevent hospitalisations and limit exposure to the virus. Several practices can reduce the chances of severe OHSS occurring. These include the use of GnRH antagonist protocol, GnRH agonist trigger, elective freeze-all and avoidance of multiple embryo transfer. As part of the closure of the trial, a survey sub-study was therefore carried out to examine if and how clinical practice at United Kingdom (UK) fertility centres changed to reduce OHSS related hospitalisations during and after the COVID-19 pandemic, to provide insight into the consistency of approaches nationally and help inform future approaches.

This paper presents data from the STOP-OHSS trial, including describing the journey of three participants randomised to receive outpatient paracentesis instead of conservative management. It also outlines the results of the post-trial survey and highlights the difficulty of conducting trials on rare conditions with rapid disease progression.

## METHODS

The STOP-OHSS trial was a pragmatic, parallel open-label, multicentre, superiority, adaptive, group sequential, confirmatory Randomised Controlled Trial. Initially designed as a 20-centre study, participants were ultimately recruited from three of the 10 National Health Service fertility centres in the UK open to recruitment. The trial protocol has been published separately^13^.

An independent Trial Steering Committee and Data Monitoring and Ethics Committee provided oversight of the trial. The trial was designed in response to a National Institute for Health Research - Health Technology Assessment Programme commissioned brief (HTA no 18/39). Ethics approval was obtained from the London - South East Research Ethics Committee (reference: 22/LO/0015) on 25^th^ February 2022. The trial was registered with ISRCTN (ISRCTN71978064). The authors of this article take accountability for the precision and completeness of the data and analyses, in addition to compliance with the protocol and interpretation of outcome results.

Regular communication with site staff occurred to ensure adherence to the trial protocol. Five minor protocol non-compliances were recorded, in relation to missed visit windows, missed visits, and missed blood tests.

### Patient and Public Involvement

Patient participation was incorporated in the delivery of the project, including in developing the funding bid and setting up study procedures. Several study documents were reviewed by the Reproductive Health Research Public Advisory Panel at Sheffield Teaching Hospitals, to improve appropriateness and clarity. There was also representation in the Trial Management Group and Trial Steering Committee. The inclusion of patients and the public was valuable to help shape the study.

### Participants

The study was designed to recruit 224 participants over 31 months. Participants were identified either through monitoring of patients with risk factors for developing moderate or severe OHSS, or through incidence cases of patients presenting with moderate or severe OHSS. Potentially eligible patients were approached after diagnosis for eligibility screening.

Patients were eligible for participation if they a) experienced moderate or severe, early or late OHSS symptoms, and b) were able and willing to attend the monitoring calls and follow-up appointments and undertake self-monitoring at home. Patients were excluded from participation if they had significant pain or vomiting requiring hospitalisation, pulmonary embolism, or their condition was judged as severe enough to warrant admission to a High Dependency Care Unit. They were also excluded if they suffered from other concurrent medical conditions requiring immediate inpatient management; if they had previously been randomised and involved in the trial but later presented again with OHSS in subsequent fertility treatment cycles; and if they were involved in other trials involving ovarian stimulation or ovarian response.

### Randomisation

Participants were randomised by the site research nurse or clinician, using a centralised web-based system (SCRAM) hosted by Sheffield Clinical Trials Research Unit (CTRU). The randomisation sequence was computer generated using random permuted blocks of sizes 2 and 4, with stratification by recruiting site and severity of OHSS. The trial statistician generated the randomisation sequence but did not have access to the generated sequence and was blinded to participant randomisation until analysis. The system had restricted access rights for delegated site staff members to enter participant details, which then generated the allocation. Participants were randomised to either receive usual care (conservative management), or outpatient paracentesis plus increased monitoring. Participants and clinicians were not blinded to their allocation due to the nature of the treatment.

### Study procedures

Following informed consent, baseline assessments were undertaken before randomisation. Both groups were asked to complete a daily diary (either on paper or received via email using Research Electronic Data Capture (REDCap)^14^). This was to collect their weight, abdominal girth, diarrhoea, vomiting, pain, shortness of breath, fluid input and urine output.

After randomisation, participants allocated to the intervention arm had outpatient paracentesis as soon as clinically possible. The study sites were recommended to adhere to their established local protocols for performing paracentesis, whether through an abdominal or vaginal approach. The procedure was carried out by an appropriately trained physician, advanced practice nurse, or radiologist from the fertility, gynaecology, or radiology department. Site staff were asked to carry out increased monitoring of these participants. This involved daily monitoring at first (either in a clinic or over the phone), which was advised to be reduced based on clinical need. The clinical team reviewed the daily diary data online (or over the phone if paper diaries) to inform further management and whether to perform further outpatient paracentesis.

Patients allocated to the control arm did not have paracentesis performed in an outpatient setting. If it was already part of their usual care, sites were asked not to conduct paracentesis in an outpatient setting for participants allocated to the control group. These patients were monitored by their clinical team as per the usual care at that site. The daily diary data was sent directly to CTRU to input to the database, for purposes of assessing the progression of the disease.

Data was also collected on the preventative measures used at the fertility centres. No other preventative measures were prohibited. Participants in both arms attended weekly monitoring visits, either face to face or remotely, at days 7, 14, 21 and 28, or until symptoms had resolved. Participants were invited to an in-person symptom resolution visit (if this did not already occur during hospitalisation) if their OHSS was resolving; indicated by normalisation of fluid input and output, and a decrease in weight and abdominal girth. Participants were also sent questionnaires at 28 days post randomisation, directly from CTRU. Additionally, data was collected from medical notes 90 days after randomisation.

### Outcomes

The primary outcome was OHSS-related hospitalisation for at least 24 hours, within 28 days of randomisation. Secondary outcomes included time to resolution of OHSS, progression of OHSS severity, cumulative length of OHSS-related hospitalisation, AEs, and participant quality of life (via the EQ-5D-5L^15^) all within 28 days of randomisation. At 28 days post-randomisation, patient satisfaction was assessed using the Client Satisfaction Questionnaire 8 (CSQ-8)^16^, and health resource use and patient costs were assessed. We captured the occurrence of thrombosis, embolism and significant infections requiring antibiotic treatment or hospitalisation within 90 days of randomisation. Finally, an independent blinded central assessment took place to adjudicate the need for OHSS-related hospitalisation within 28 days.

### Statistical methods

The trial initially intended to report the unadjusted difference in hospitalisation rates with confidence intervals obtained via normal approximation, as well as the p-value from x^2^ test. Details on this, and analyses planned for the secondary outcomes, can be found in the protocol paper^13^. Due to the low recruitment, such analyses were not possible, therefore we have presented participant-level data with some summary statistics.

### Closure survey

After all 90-day follow-ups were completed, a survey was sent to fertility centres in the UK. Questions related to the practices that centres carry out, and whether these changed during and after the COVID-19 pandemic. Questions were mostly closed with multiple choice answers, and there were a small number of open questions with a free text box to provide more detail. The survey questions can be found here (link). Respondents were encouraged, but not required, to answer all questions. Ethical review was not required, as the survey did not involve any patient data collection or transfer of any confidential information to researchers.

Throughout the survey we referred to before/during/after COVID-19, highlighting that the World Health Organisation declared the outbreak as a Public Health Emergency of International Concern (PHEIC) on 30th January 2020, then no longer a PHEIC on 5th May 2023.

Responses were sought from as many fertility centres as possible in the UK, with emails sent to all fertility centres in the UK for which the team could obtain contact details. The survey was completed by one member of staff at each site (e.g., a clinician, embryologist, centre manager, or research nurse).

The survey was hosted on the online Qualtrics platform Version January 2023^17^. All fertility centres that completed the survey were entered into a prize draw for a chance to win one of five food delivery vouchers for the local team. Data was exported from Qualtrics and saved in a locked Excel file. Data was analysed to produce summary statistics. All data were anonymised before analysis.

## RESULTS

### Participant flow and recruitment

A total of 146 patients were screened. Of these, 137 (93.8%) were excluded (Figure 1), predominantly for not being eligible (n=137). Nine patients consented to be randomised, but the randomisation window was missed for one patient due to delayed blood test results. Eight participants were therefore randomised between 5^th^ November 2022 and 25^th^ May 2023. Five were allocated to receive conservative management (CM), and three to receive outpatient paracentesis (OP). Figure 1 shows the flow of participants during their follow-up on days 7, 14, 21, and 28 as well as day 90 safety monitoring via medical records.

**Figure 1.**
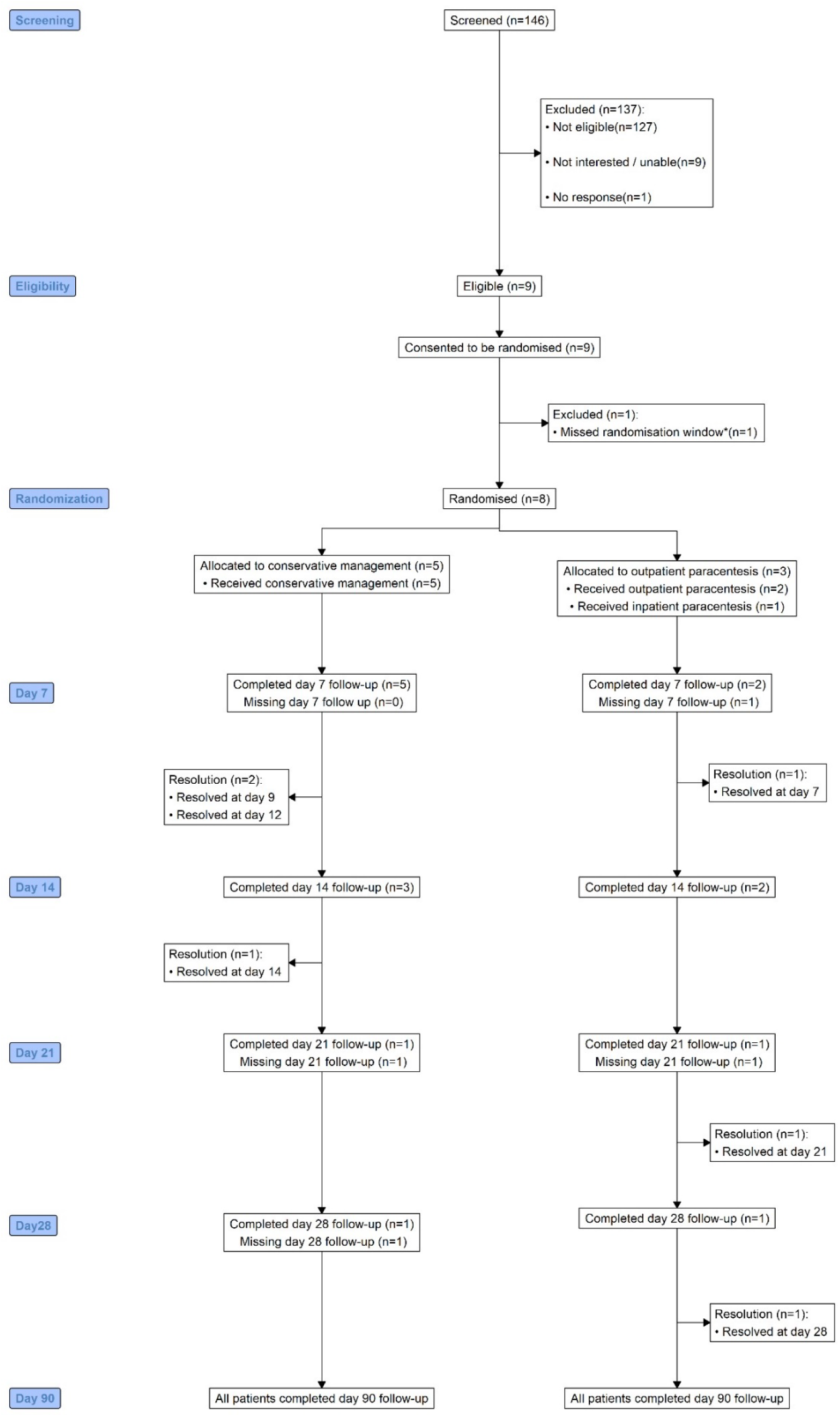
Flow of participants.

### Demographics and baseline data

Participants were recruited from three of the nine sites that screened patients for eligibility. Participants’ age at consent ranged from 26 to 42 years. Only one participant in the CM self-identified as Indian and the rest were English/ Welsh/ Scottish/ Northern Irish/ British. All eight participants had moderate OHSS; five had early OHSS and three had late OHSS.

Seven participants were on antagonist fertility treatment. Three participants in the CM arm had been prescribed Cabergoline medication for OHSS. Four participants (two in each arm) had previously been diagnosed with polycystic ovary syndrome. Two participants (one in each arm) had a history of moderate OHSS as their highest experienced severity. Demographic information is shown in Table 1.

**Table 1.** Demographics and baseline characteristics of 8 randomised participants.

|  | **Conservative**<br>**management (n=5)** | **Outpatient**<br>**paracentesis (n=3)** | **Total (n=8)** |
| --- | --- | --- | --- |
| **Ethnicity** |  |  |  |
| English/Welsh/Scottish/Northern Irish/British | 4 (80.0%) | 3 (100.0%) | 7 (87.5%) |
| Indian | 1 (20.0%) | 0 (0.0%) | 1 (12.5%) |
| **OHSS classification** |  |  |  |
| Early | 3 (60.0%) | 2 (66.7%) | 5 (62.5%) |
| Late | 2 (40.0%) | 1 (33.3%) | 3 (37.5%) |
| **Fertility treatment** |  |  |  |
| Antagonist protocol | 4 (80.0%) | 3 (100.0%) | 7 (87.5%) |
| Short (flare) protocol | 1 (20.0%) | 0 (0.0%) | 1 (12.5%) |
| Cabergoline prescribed | 3 (60.0%) | 0 (0.0%) | 3 (37.5%) |
| History of PCOS diagnosis | 2 (40.0%) | 2 (66.7%) | 4 (50.0%) |
| Previous moderate OHSS | 1 (20.0%) | 1 (33.3%) | 2 (25.0%) |
| Mean age at consent (years) | 32.0 | 28.0 | 31.5 |
| Mean body mass index (BMI, kg/m<sup>2</sup>) | 26.7 | 27.4 | 26.7 |
| Mean weight (kg) | 74.2 | 62.9 | 69.7 |
| Mean height (cm) | 162.6 | 152.4 | 162.6 |
| Mean abdominal girth (cm) | 100.0 | 86.4 | 99.3 |

Presentation of continuous variables: median (minimum - maximum); \* 2 of the 3 had available data; otherwise, the denominator was the same as randomised for all the continuous outcomes. PCOS = Polycystic Ovary Syndrome; BMI = Body Mass Index.

### OHSS hospitalisation, treatment and progression

Two participants were hospitalised for OHSS-related reasons, agreed by the independent adjudication panel: one had been randomised to CM, and the other to OP. Both had late OHSS.

The hospitalised participant in the OP arm was admitted for approximately 246 hours due to an increase in symptom severity: abdominal pain and swelling, nausea, shortness of breath, vomiting and moderate pleural effusion, and constipation. This patient received inpatient paracentesis as an abdominal procedure on day five following randomisation, which resulted in 135ml of ascitic fluid removed.

The two other participants randomised to OP received OP as the first and only procedure (Table 2). Both underwent the procedure using the vaginal technique – one on the same day as randomisation, with 200ml of fluid removed, and the other on the second day following randomisation, with 40ml removed.

**Table 2.** Characterisation of paracentesis in participants randomised to OP.

| Randomised treatment | Paracentesis location | Time to procedure from randomisation (days) | Technique | Clinician performing | Volume of ascites removed (ml) | IV fluids given | IV colloid therapy given |
| --- | --- | --- | --- | --- | --- | --- | --- |
| OP | Outpatient | 0 | Transvaginal | Doctor | 200 | No | No |
| OP | Outpatient | 2 | Transvaginal | Doctor | 40 | No | No |
| OP | Inpatient | 5 | Abdominal | Doctor | 135 | No | No |

OP, outpatient paracentesis. CM, conservative management. IV, intravenous.

The hospitalised participant in the CM group was admitted for approximately 89.3 hours. This patient had received Cabergoline beforehand. They were hospitalised for rehydration, with concerns of OHSS progression, however, they remained at a moderate diagnosis. This participant received inpatient paracentesis during this hospitalisation, which was performed on day two of randomisation. This was performed transvaginally, and 3000ml of ascitic fluid was removed. The two other participants randomised to CM only received medications for OHSS.

No participants were prescribed a GnRH antagonist during the trial. Four of the eight participants (two in CM and two in OP) were given prescribed or concomitant medications after randomisation. Details of these medications including associated indication, dose, frequency and route of administration are presented in Appendix 1.

All participants had moderate OHSS at baseline and their OHSS severity did not deteriorate compared to their baseline state. Most patients’ symptoms resolved during the follow-up period. One patient in the CM arm missed the day 21 and 28 follow-up and one patient in the OP arm missed the day 21 follow-up. The OHSS severity of six participants decreased from moderate to mild with no progression thereafter. The OHSS severity of one participant in the OP group fluctuated initially, dropping to mild briefly before returning to moderate, and finally stabilizing at mild by the end of the follow-up period (Figure 2). Seven participants had daily diary data relating to fluid input and output, abdominal girth, and body weight; this data is presented in Appendix 2.

**Figure 2.**
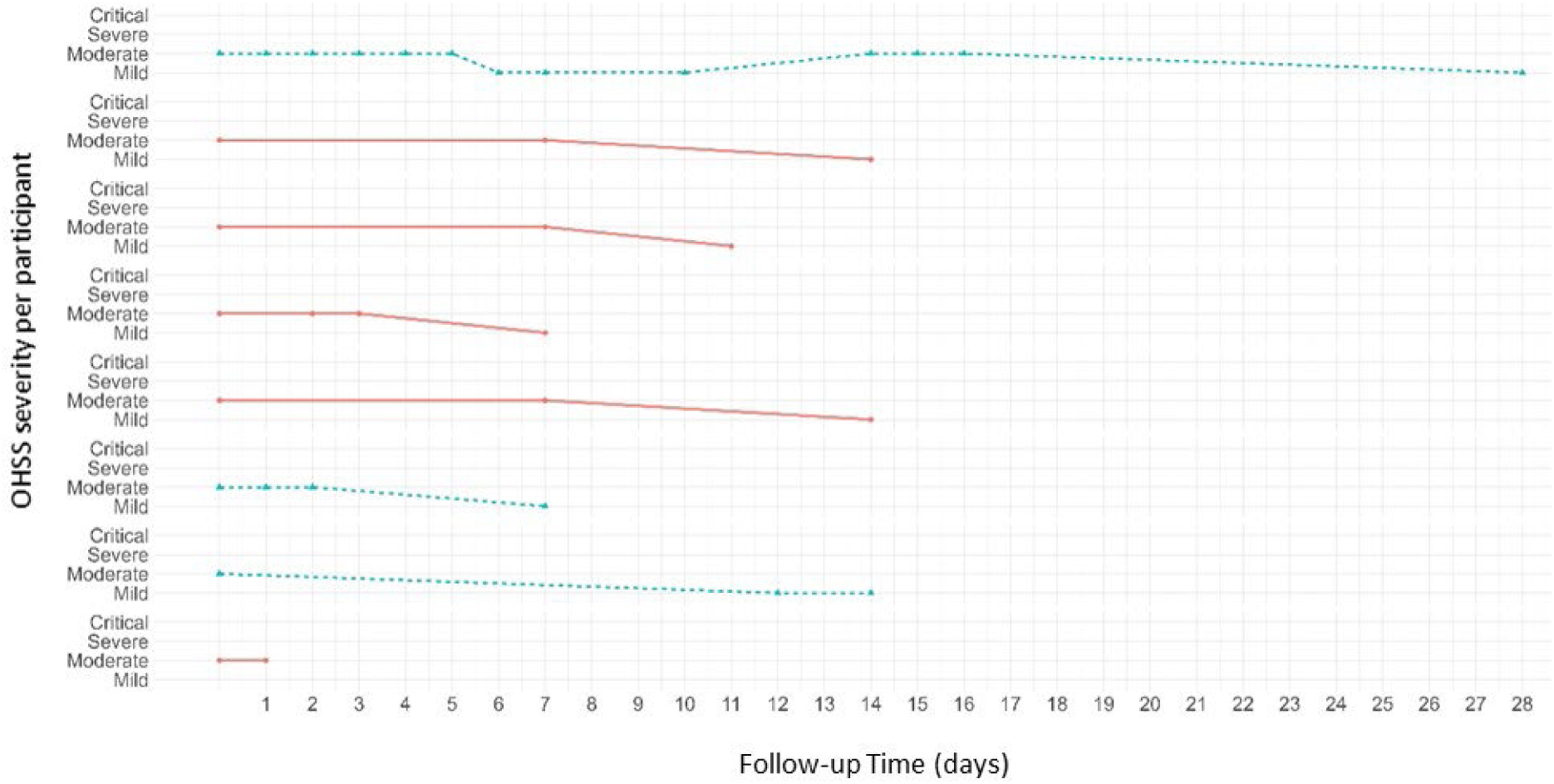
Profile of OHSS severity over time for the eight individual participants.

### Patient satisfaction

Only half of the participants (three in CM vs one in OP) completed the CSQ-8^16^ at 28 days post-randomisation. One participant, from the CM group, reported a low satisfaction score and the rest were extremely satisfied (Table 3).

**Table 3.** Listing of treatment characteristics and client satisfaction scores on day 28.

| Treatment group | Received paracentesis | Location | CSQ 8 total score |
| --- | --- | --- | --- |
| OP | Yes | Outpatient | 31 |
| CM | No | n/a | 32 |
| CM | No | n/a | 15 |
| CM | No | n/a | 32 |

CSQ-8 = client satisfaction questionnaire 8. The higher the score the better the client satisfaction.

### Harms

A total of six adverse events (including repeated events) were reported. Three occurred in the same participant randomised to OP: hyperemesis, very slight vaginal bleeding, and per vaginal bleeding. The hyperemesis was reported as an unexpected Serious Adverse Event of moderate intensity, but was assessed as caused by pregnancy, not related to the OP. One participant randomised to CM presented with low mood, and another had symptoms of a urinary tract infection, though no infection was found. All 8 participants had data at day 90. No participants experienced thrombosis or embolism within 90 days of randomisation and one participant who was randomised to OP reported an infection that did not require hospitalisation on day 90. There were no other Serious Adverse Events. Details of the adverse events can be found in Appendix 3.

### Post-trial Survey

Responses were received between 31/10/2024 and 22/01/2024, from a total of 16 fertility centres (Table 4) across the UK. Ten of these centres were already open to the main STOP-OHSS trial, and six were other centres. The data obtained can be broken down into general descriptive data on fertility practices, and changes due to the COVID-19 pandemic.

**Table 4.** Site (n = 16) responses to the post-trial survey on clinic practice.

| Question | Site response (n=16) |  |
| --- | --- | --- |
|  | Yes | No |
| Do you routinely undertake planned “freeze all” cycles (not including fertility preservation for medical reasons)? | 13 | 5 |
| Has your elective freeze rate for prevention of OHSS changed during/after COVID-19? | 10 | 6 |
| Is your clinical pregnancy rate (per egg transfer) from frozen embryos the same as it is for fresh embryos? | 8 | 7 |
| Did the e-freeze study influence your practice of freezing? | 1 | 13 |
| Do you routinely reduce the dose of stimulating hormones (drugs) during monitoring if you find an excessive ovarian response? | 11 | 5 |
| Do you use an antagonist protocol? | 16 | 0 |
| Did your practice of this change during COVID-19? (increased) | 8 | 7 |
| Has this change been maintained after COVID-19? | 6 | 2 |
| Do you use cabergoline for prevention of OHSS? | 8 | 8 |
| Did your practice of this change during COVID-19? (increased) | 1 | 7 |
| Has this change been maintained after COVID-19? | 1 | 0 |
| Do you use GnRH trigger rather than hCG in women at risk of OHSS? | 16 | 0 |
| Did your practice of this change during COVID-19? (increased) | 8 | 7 |
| Has this change been maintained after COVID-19? | 8 | 0 |
| Did you change the dose of FSH in your standard protocols during COVID-19? | 0 | 16 |
| Did your single embryo transfer rate change during COVID-19? (increased) | 2 | 14 |
| Has this change in practice been maintained since COVID-19? | 1 | 1 |
| Did your single embryo transfer rate change during COVID-19? (decreased) | 1 | 15 |
| Has this change in practice been maintained since COVID-19? | 0 | 0 |
| Does your unit carry out paracentesis in the outpatient setting? | 3 | 12 |
| Is this a practice you are considering in the future? | 7 | 3 |

OHSS = Ovarian Hyperstimulation Syndrome; GnRH = Gonadotropin-Releasing Hormone; FSH = Follicle-Stimulating Hormone;

### Descriptive data

Where sites responded to a question with ‘unknown’, the response has been excluded from the percentages in this section. 81% (n = 13) of sites reported that they routinely undertake freeze-all cycles. 50% (n = 8) stated that their clinical pregnancy rate is the same for frozen and fresh embryos. 47% (n = 7) of sites reported that they do freeze embryos at the cleavage/pronucleate stage, with a post-thaw survival rate of on average 87.2%. 69% (n = 11) routinely reduce the dose of stimulating hormones during monitoring if an excessive ovarian response is found. All 16 sites reported that they use an antagonist protocol, and likewise, all 16 sites use a GnRH trigger rather than an hCG trigger in women at risk of OHSS. 50% (n = 8) use cabergoline for prevention of OHSS.

At the time of completing the survey, three sites stated that they currently carry out paracentesis in the outpatient setting. Seven were considering implementing this in the future, three were not considering this, and two did not know.

### Changes due to the COVID-19 pandemic

31% (n = 5) of respondents believed that the incidence of moderate and severe OHSS had decreased during and after the pandemic, whereas the remaining 11 respondents believed it did not change. However, a number of changes in preventive practices were reported to be made during the pandemic, and maintained after. Additionally, 91% (n = 10) of sites that routinely undertake freeze-all cycles increased their elective freeze rate from before to post pandemic, by an increase ranging from 0.6% to 20% (See Table 5). One further site increased their elective freeze rate during the pandemic but did not maintain this after.

**Table 5.**
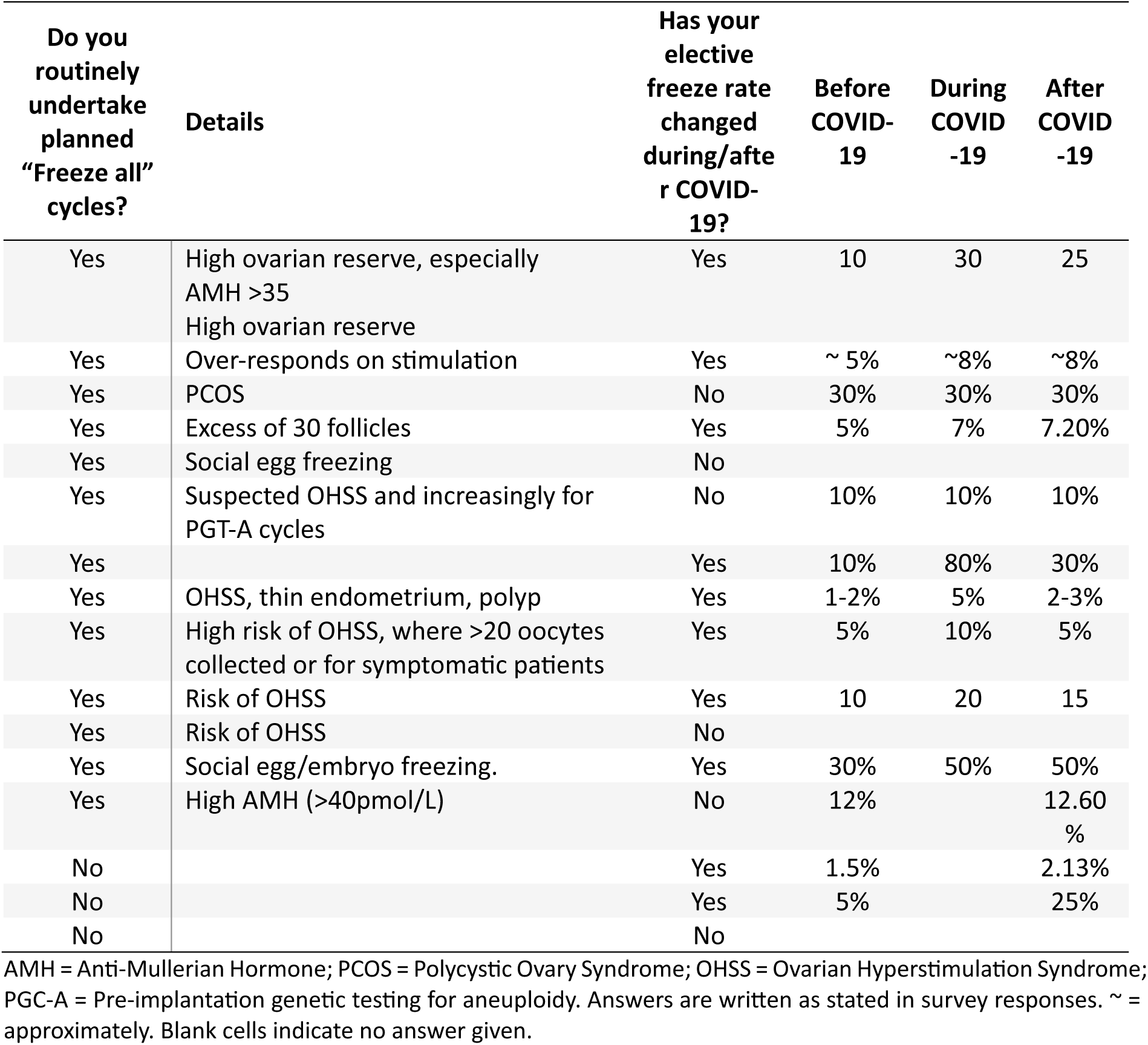
Approximate freeze rates of sites before, during and after COVID-19, reported in the post-trial survey.

53% (n = 8) of sites reported that they increased their use of the antagonist protocol during the pandemic, and of these, 75% (n = 6) maintained this increase after. 53% (n = 8) of sites increased their use of GnRH trigger and maintained this increase after the pandemic. 13% (n = 1) of the 8 sites that use cabergoline increased their use of cabergoline during and after the pandemic. 14% (n = 2) increased their single embryo transfer rate, and one of these sites maintained this increase, but the other did not know if it was maintained. 7% (n = 2) decreased their single embryo transfer rate, but did not maintain this after the pandemic. Finally, no sites changed the dose of Follicle-Stimulating Hormone in their standard protocols during the COVID-19 pandemic.

## DISCUSSION

Given the limited sample size of this study, the findings presented are primarily descriptive in nature, precluding the ability to draw definitive conclusions regarding the comparative effectiveness and safety of outpatient paracentesis versus conservative management approaches. The planned cost-effectiveness analysis was not undertaken. While the intervention groups appear to be broadly comparable in terms of baseline characteristics, the small number of participants in this study significantly limits the ability to derive statistically meaningful comparisons or draw conclusions. Nevertheless, the analysis of these eight cases of moderate ovarian hyperstimulation syndrome (OHSS) suggests that outpatient paracentesis may be a promising and safe option for managing OHSS patients. Very low incidence rates of OHSS were observed across UK sites. Though the Human Fertilisation and Embryology Authority publish information on the reported cases of OHSS, it is difficult to interpret trends over the past few years for two main reasons. In 2017 a newspaper alleged under-reporting of clinics of OHSS^18^. As a result, a new reporting form was introduced in mid-2018. Subsequently in 2021/2022 clinics were following recommendations to report all hospitalisations due to the pandemic, leading to more mild and moderate cases of OHSS being reported^11,19,20^. It is therefore not possible to infer from this data whether the OHSS incidence was affected by the pandemic. Our post-trial survey provides some insight into this. Approximately one-third of respondents believed that there had been a decrease in moderate and severe OHSS incidence after COVID-19; although most sites reported that there had been no change in incidence, they did report an increase in preventative strategies. This included an increase in elective freeze-all cycles in almost all sites, and a greater use of antagonist protocols and GnRH trigger in approximately half of the sites. It should be noted that survey responses were received from 16 of 107 Human Fertilisation and Embryology Authority -licensed fertility centres across the UK (in 2022/2023)^21^ and so may not be fully representative of UK practice. Additionally, responses were the subjective opinion of the member of staff who completed the survey.

The effectiveness of outpatient paracentesis in this population remains unclear. Our post-trial survey did reveal that three sites were already carrying out paracentesis in the outpatient setting, and a further seven were thinking of implementing this in the future, therefore this may be a practice which will increase in the coming years. Although further randomised controlled trials to assess this with a larger sample size would be ideal, the low incidence of OHSS and the challenges of recruiting this population to a trial raise questions about the feasibility of future definitive randomised trials - trials on rare conditions, particularly those with rapid disease progression, can be difficult to recruit to. This is evidenced by the fact that randomisation was missed for one participant who consented, as they presented out of routine clinic hours and the site could not reach the research team to discuss the delayed blood test results, despite the research teams best efforts to develop study management plans facilitating recruitment out of hours. This resulted in ultimately missing the window before the patient’s OHSS resolved and they became ineligible. Researchers should consider innovative study designs and collaborative efforts to overcome these inherent challenges.

We encountered difficulties in attempting to adjudicate the time it took for symptoms to resolve. This assessment was based on participants’ self-reported diary entries, which included data on fluid intake and output, abdominal circumference, and weight. There was considerable variation in how different adjudicators interpreted the data, making it impossible to determine a consistent timeline for symptom resolution. The date of the ‘symptom resolution visit’ organised by the site was therefore considered to be their date of resolution.

In conclusion, this study provides preliminary insights into the use of outpatient paracentesis for managing moderate OHSS, despite limitations due to its small sample size. While the findings suggest that outpatient paracentesis may be a promising and safe option, definitive conclusions about its effectiveness compared to conservative management cannot be drawn. The study highlights significant challenges in researching OHSS, which may impede future large-scale randomised trials. Our post-trial survey revealed evolving practices in OHSS prevention and management across UK sites.

## AUTHOR CONTRIBUTIONS

Conceptualisation: Ying Cheong, Munya Dimairo, Andrew Drakeley, Raj Mathur, Mostafa Metwally, Alicia O’Cathain, Clare Pye, David White. Data curation: Munya Dimairo, Amanda Loban, Katie Ridsdale, Rich Simmonds, Siqi Wu. Formal analysis: Lauren Desoysa, Munya Dimairo, Siqi Wu. Funding acquisition: Ying Cheong, Munya Dimairo, Andrew Drakeley, Raj Mathur, Mostafa Metwally, Clare Pye, David White. Investigation: Ying Cheong, Andrew Drakeley, Isaac Evbuomwan, Raj Mathur, Mostafa Metwally, Clare Pye, Liz Taylor. Methodology: Ying Cheong, Munya Dimairo, Andrew Drakeley, Isaac Evbuomwan, Raj Mathur, Mostafa Metwally, Cara Mooney, Clare Pye, David White. Project administration: Jane Hughes, Kirsty McKendrick, Mostafa Metwally, Cara Mooney, Clare Pye, Katie Ridsdale, David White. Software: Lauren Desoysa, Munya Dimairo, Amanda Loban, Rich Simmonds, Siqi Wu. Supervision: Munya Dimairo, Mostafa Metwally, Alicia O’Cathain, David White. Validation: Munya Dimairo. Visualisation: Munya Dimairo, Katie Ridsdale, David White, Siqi Wu. Writing – original draft: Munya Dimairo, Mostafa Metwally, Katie Ridsdale, David White. Writing – review & editing: All.

## DATA SHARING STATEMENT

All data requests should be submitted to the corresponding author for consideration. De-identified participant data and statistical code will be made available upon reasonable request. Requests should state the data fields required and purpose of the request (ideally with a protocol but, at a minimum, with a research plan). The data dictionary and statistical analysis plan can also be made available. Requests will be considered on a case-by-case basis and requestors will be asked to complete a data sharing agreement with the sponsor before data transfer.

## Appendix 1. Concomitant medications

Prescribed medication and concomitant medications to five participants after randomisation

| Treatment group | Paracentesis Location | Medication | Specific medication | Indication | Dose | Unit | Frequency | Route | Start date | End date |
| --- | --- | --- | --- | --- | --- | --- | --- | --- | --- | --- |
| OP | Outpatient | Other | Paracetamol | Pain relief | 1 | g | PRN | PO | 2022-12-19 | 2023-01-16 |
| OP | Outpatient | Other | Prochlorperazine | Antiemetic | 20 | mg | PRN | PO | 2023-01-05 | 2023-01-16 |
| OP | Outpatient | Other | Cyclizine | Antiemetic | 50 | mg | PRN | PO | 2023-01-02 | 2023-01-16 |
| OP | Outpatient | Other | Dalteparin | Anticoagulant | 5000 | IU | OD | SC | 2022-12-19 | 2023-01-16 |
| OP | Outpatient | Other | Fentanyl | Pain relief | 75 | µg | STAT | IV | 2022-12-19 | 2022-12-19 |
| OP | Outpatient | Other | Paracetamol | Pain relief | 1 | g | STAT | IV | 2022-12-19 | 2022-12-19 |
| OP | Outpatient | Other | Ondansetron | Antiemetic | 4 | mg | STAT | IV | 2022-12-19 | 2022-12-19 |
| OP | Outpatient | Other | Cyclizine | Antiemetic | 50 | mg | STAT | IM | 2022-12-19 | 2022-12-19 |
| CM | n/a | Other | Dalteparin | Anticoagulant | 5000 | IU | OD | SC | 2022-12-22 | 2023-01-19 |
| CM | n/a | Other | Paracetamol | Pain relief | 1 | g | PRN | PO | 2022-12-22 | 2023-01-12 |
| CM | Inpatient | Other | Fragmin | Thromboprophylaxis | 5000 | IU | OD | SC | 2022-11-05 | 2023-01-07 |
| CM | Inpatient | Other | Paracetamol | Pain relief | 1 | g | OD | IV | 2022-11-06 | 2022-11-09 |
| CM | Inpatient | Other | Normal Saline | Fluids | 1 | % | OD | IV | 2022-11-06 | 2022-11-07 |
| OP | Inpatient | Other | Ondansetron | Nausea | 4 | mg | TDS | PO | 2023-05-13 |  |

OP, patient paracentesis; g, gram, mg, milligram, µg, microgram; IU, international unit; PRN, as required; OD, once daily; STAT, once only; TDS, three times a day; PO, oral; SC, subcutaneous; IV, intravenous; IM, intramuscular

## Appendix 2. Daily diary data.

Seven of the eight participants had daily diary data relating to fluid input and output (from which fluid balance was derived), abdominal girth, and body weight. The figures below display seven participants’ profiles of these symptoms over 28 days after randomisation stratified by randomised group. Some gaps exist between periods due to missing day(s) of diary data.

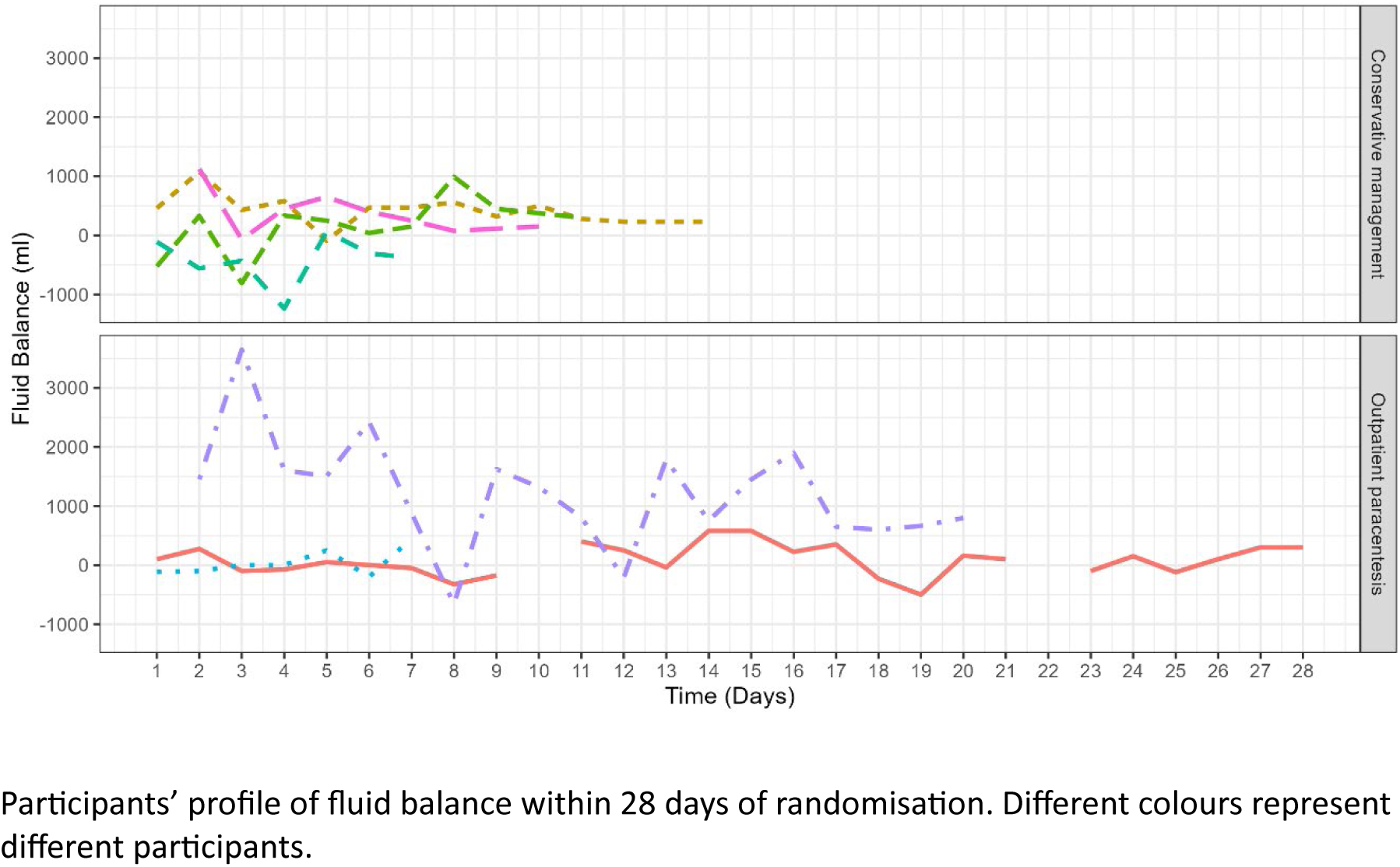

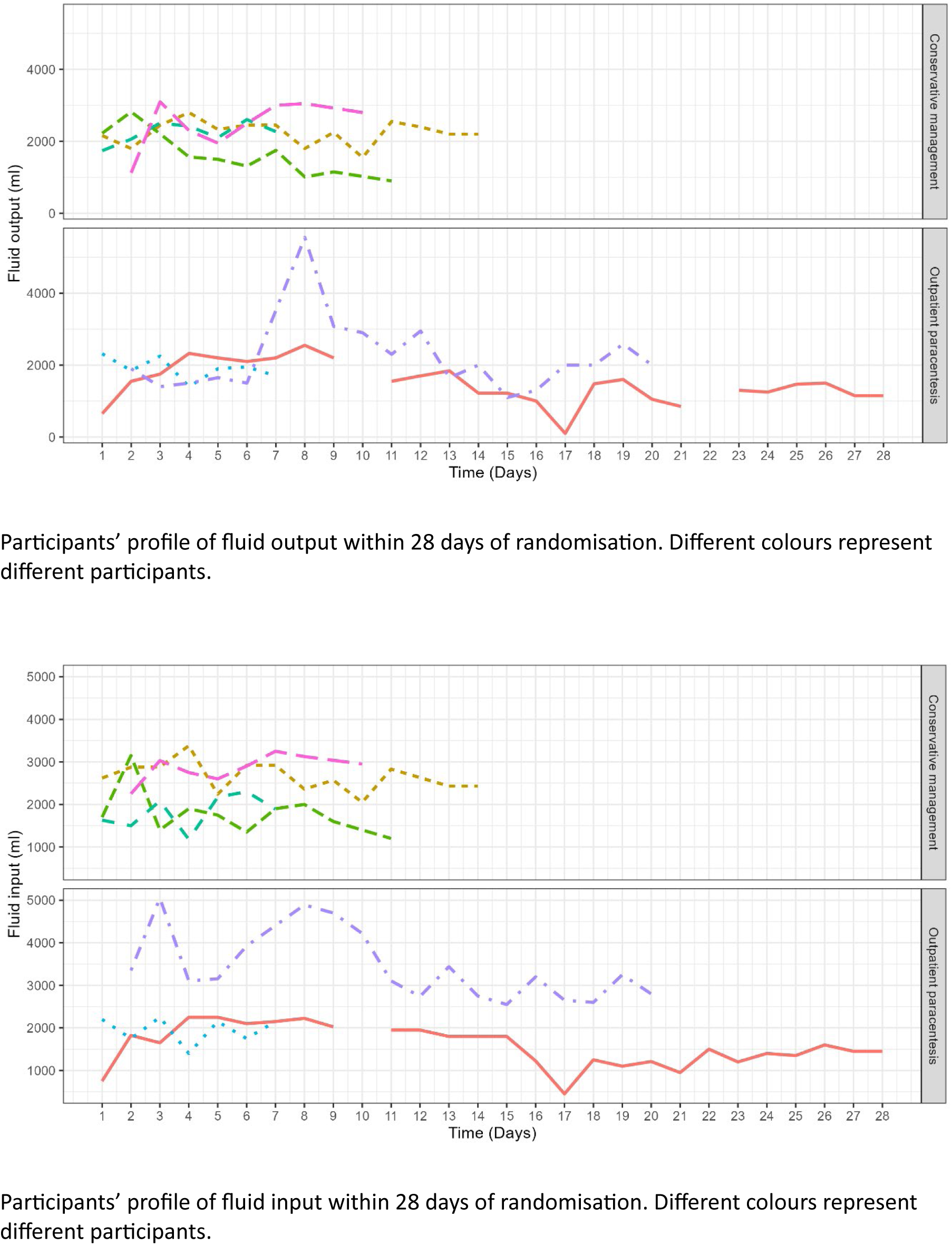

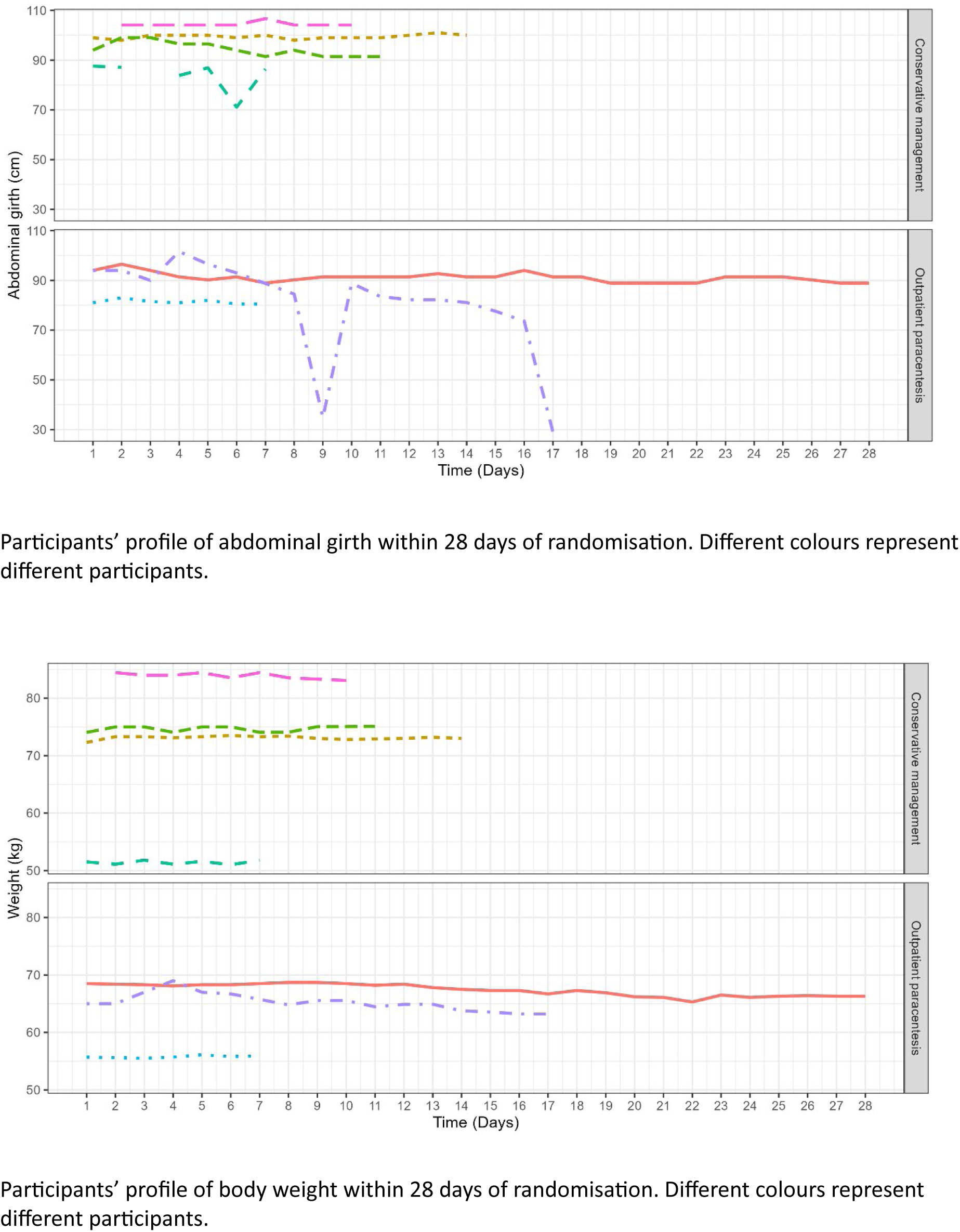

## Appendix 3. Adverse events.

Adverse and serious adverse events as per randomised treatment.

| Adverse event classification | Conservative management |  | Outpatient paracentesis |  |
| --- | --- | --- | --- | --- |
|  | (N = 5)<sup>a</sup> | n<sup>b</sup> | (N = 3)<sup>a</sup> | n<sup>b</sup> |
| AEs | 2(40.0%) | 2 | 2(66.7%) | 4 |
| SAEs | 0(0.0%) | 0 | 1(33.3%) | 1 |
| Unexpected SAEs | 0(0.0%) | 0 | 1(33.3%) | 1 |
| Expected SAEs | 0(0.0%) | 0 | 0(0.0%) | 0 |
| Details of the SAE category |  |  |  |  |
| Seriousness |  |  |  |  |
| Inpatient hospitalisation | 0(0.0%) | 0 | 1(33.3%) | 1 |
| Frequency |  |  |  |  |
| Isolated | 0(0.0%) | 0 | 1(33.3%) | 1 |
| Intensity |  |  |  |  |
| Moderate | 0(0.0%) | 0 | 1(33.3%) | 1 |
| Outcome |  |  |  |  |
| Recovered | 0(0.0%) | 0 | 1(33.3%) | 3 |
| Relationship to OP |  |  |  |  |
| No reasonable possibility of being related | 0(0.0%) | 0 | 1(33.3%) | 1 |

<sup>a</sup> column statistics relates to the number and proportion of participants with at least one event in a specific category; <sup>b</sup> n relates to the number of all events including repeated events per participant; OP, outpatient paracentesis; AE, adverse event; SAE, serious adverse event.

Adverse events and serious adverse events as per treatment received.

| Adverse events classification | Conservative management |  | Outpatient paracentesis |  |
| --- | --- | --- | --- | --- |
|  | (N = 6)<sup>a</sup> | n<sup>b</sup> | (N = 2)<sup>a</sup> | n<sup>b</sup> |
| AEs | 3(50.0%) | 3 | 1(50.0%) | 3 |
| SAEs | 0(0.0%) | 0 | 1(50.0%) | 1 |
| Unexpected SAEs | 0(0.0%) | 0 | 1(50.0%) | 1 |
| Expected SAEs | 0(0.0%) | 0 | 0(0.0%) | 0 |
| Details of the SAE category |  |  |  |  |
| Seriousness |  |  |  |  |
| Inpatient hospitalisation | 0(0.0%) | 0 | 1(50.0%) | 1 |
| Frequency |  |  |  |  |
| Isolated | 0(0.0%) | 0 | 1(50.0%) | 1 |
| Intensity |  |  |  |  |
| Moderate | 0(0.0%) | 0 | 1(50.0%) | 1 |
| Outcome |  |  |  |  |
| Recovered | 0(0.0%) | 0 | 1(50.0%) | 3 |
| Relationship to OP |  |  |  |  |
| No reasonable possibility of<br>being related | 0(0.0%) | 0 | 1(50.0%) | 1 |

<sup>a</sup> column statistics relates to the number and proportion of participants with at least one event in a specific category; <sup>b</sup> *n* relates to the number of all events including repeated events per participant; OP, outpatient paracentesis; AE, adverse event; SAE, serious adverse event.

## Appendix 4. The STOP-OHSS Study Group

Amy Barr, Sarah Drabble, Laura Flight, Anju Keetharuth, Elizabeth Lumley, Alicia O’Cathain, Jessica Wright, Tracey Young.

## REFERENCES

1. Royal College of Obstetricians and Gynaecologists. RCOG Green-top Guideline No. 5: The Management of Ovarian Hyperstimulation Syndrome [Online]. 2016. https://www.rcog.org.uk/guidance/browse-all-guidance/green-top-guidelines/the-management-of-ovarian-hyperstimulation-syndrome-green-top-guideline-no-5/ (accessed 25 June 2024).

2. Smith LP, Hacker MR, Alper MM. Patients with severe ovarian hyperstimulation syndrome can be managed safely with aggressive outpatient transvaginal paracentesis. Fertil Steril. 2009 Dec;92(6):1953–9. doi: 10.1016/j.fertnstert.2008.09.011

3. Lincoln SR, Opsahl MS, Blauer KL, Black SH, Schulman JD. Aggressive outpatient treatment of Ovarian Hyperstimulation Syndrome with ascites using transvaginal culdocentesis and intravenous albumin minimizes hospitalization. J Assist Reprod Genet. 2002;19(4):159–63. doi: 10.1023/A:1014828027282

4. Gustofson RL, Browne P, Van Nest RL, Richter KS, Larsen FW. Aggressive Outpatient Management of Severe Ovarian Hyperstimulation Syndrome Avoids Complications and Prolonged Disease Course. Fertil Steril. 2005 Sep;84:S95. doi: 10.1016/j.fertnstert.2005.07.229

5. Fluker MR, Copeland JE, Yuzpe AA. An ounce of prevention: Outpatient management of the ovarian hyperstimulation syndrome. Fertil Steril. 2000 Apr;73(4):821–4. doi: 10.1016/S0015-0282(99)00606-8

6. Shrivastav P, Nadkarni P, Craft I. Day care management of severe ovarian hyperstimulation syndrome avoids hospitalization and morbidity. Hum Reprod. 1994 May;9(5):812–4. doi: 10.1093/oxfordjournals.humrep.a138601

7. Abuzeid M, Warda H, Joseph S, Corrado MG, Abuzeid Y, Ashraf M, et al. Outpatient Management of Severe Ovarian Hyperstimulation Syndrome (OHSS) with Placement of Pigtail Catheter. Facts, Views Vis ObGyn. 2014;6(1):31. doi: 10.4274/tjod.33340

8. Abuzeid MI, Nassar Z, Massaad Z, Weiss M, Ashraf M, Fakih M. Pigtail catheter for the treatment of ascites associated with ovarian hyperstimulation syndrome. Hum Reprod. 2003 Feb;18(2):370–3. doi: 10.1093/humrep/deg074

9. Gul RB, Ali PA. Clinical trials: the challenge of recruitment and retention of participants. J Clin Nurs. 2010 Jan;19(1–2):227–33. doi: 10.1111/J.1365-2702.2009.03041.X

10. Mitchell EJ, Ahmed K, Breeman S, Cotton S, Constable L, Ferry G, et al. It is unprecedented: trial management during the COVID-19 pandemic and beyond. Trials 2020 21:1. 2020 Sep 11;21(1):1–7. doi: 10.1186/S13063-020-04711-6

11. HFEA. Research and data [online]. https://www.hfea.gov.uk/about-us/publications/research-and-data (accessed 29 July 2024)

12. Lumley E, O’Cathain A, Drabble S, Pye C, Brian K, Metwally M. Managing ovarian hyperstimulation syndrome: A qualitative interview study with women and healthcare professionals. J Clin Nurs. 2023;32(17–18):6599–610. doi: 10.1186/S13063-020-04711-6

13. White DA, Pye C, Ridsdale K, Dimairo M, Mooney C, Wright J, et al. Outpatient paracentesis for the management of ovarian hyperstimulation syndrome: study protocol for the STOP-OHSS randomised controlled trial. BMJ Open. 2024;14(1):1–11. doi: 10.1136/bmjopen-2023-076434

14. Harris PA, Taylor R, Thielke R, Payne J, Gonzalez N, Conde JG. Research electronic data capture (REDCap)—A metadata-driven methodology and workflow process for providing translational research informatics support. J Biomed Inform. 2009 Apr;42(2):377–81. doi: 10.1016/J.JBI.2008.08.010

15. Van Hout B, Janssen MF, Feng YS, Kohlmann T, Busschbach J, Golicki D, et al. Interim scoring for the EQ-5D-5L: Mapping the EQ-5D-5L to EQ-5D-3L value sets. Value Heal. 2012 Jul;15(5):708–15. doi: 10.1016/j.jval.2012.02.008

16. Larsen DL, Attkisson CC, Hargreaves WA, Nguyen TD. Assessment of client/patient satisfaction: Development of a general scale. Eval Program Plann. 1979;2(3):197–207. doi: 10.1016/0149-7189(79)90094-6

17. Qualtrics. Qualtrics XM [online]. 2005. https://www.qualtrics.com/ (accessed 29 July 2024)

18. Fertility clinics accused of covering up IVF side effects [online]. Daily Mail Online. https://www.dailymail.co.uk/news/article-4471726/Fertility-clinics-accused-covering-IVF-effects.html (accessed 29 July 2024)

19. HFEA. Impact of COVID-19 on fertility treatment 2020 (online). https://www.hfea.gov.uk/about-us/publications/research-and-data/impact-of-covid-19-on-fertility-treatment-2020/ (accessed 29 July 2024)

20. Verdin H. Ovarian hyperstimulation syndrome. Human Fertilisation and Embryology Authority [online]. 2018. http://qna.files.parliament.uk/qna-attachments/1012110/original/2018-01-24 - Authority paper - item 10 - Ovarian hyperstimulation syndrome - FINAL.doc (accessed 29 July 2024).

21. HFEA. State of the fertility sector 2022/23 (online). https://www.hfea.gov.uk/about-us/publications/research-and-data/state-of-the-fertility-sector-2022-2023/ (accessed 19 July 2024)

